# Performance of a Supervised Nurse-led Outpatient Clinic for Difficult-to-manage Gout Patients at a Tertiary Center

**DOI:** 10.64898/2026.01.08.26343694

**Authors:** Piroska Maurer, Regina Herren, Tobias Manigold

**Affiliations:** Faculty of Medicine, University of Bern, Switzerland; University Clinic for Rheumatology and Immunology, University Hospital Bern, Switzerland; Department of Rheumatology, Kantonsspital Baden, Switzerland

**Keywords:** Gout, Nurse-led management, Difficult-to-treat gout, Tertiary care, Transplant patients

## Abstract

**Background:** Gout is the among the most prevalent arthritides worldwide and is associated with high morbidity, cardiovascular risk, and substantial healthcare burden. Despite effective urate-lowering therapies (ULT), many patients remain undertreated, particularly those with severe or complex disease. Nurse-led gout management has improved outcomes in primary care, but data in multimorbid tertiary center populations are lacking.

**Objective:** To evaluate the effectiveness of a supervised nurse-led gout clinic in achieving and maintaining serum urate (UA) targets in a tertiary hospital cohort.

**Methods:** In this observational study at University Hospital Bern (May 2023–Jan 2026), 69 patients with confirmed gout were enrolled. Patients were assigned to UA targets of <300 µmol/L (Group 1: severe gout) or <360 µmol/L (Group 2: non-severe gout). An advanced Practice Nurse (APN) provided education, a rheumatologist provided pharmacological management per 2020 ACR guidelines. UA levels, flare frequency, prophylaxis, and treatment adjustments were monitored, with follow-up at six and twelve months.

**Results:** Group 1 required higher allopurinol doses than Group 2 (p<0.01). UA targets were achieved in 82% of Group 1 regimens and 68% of Group 2. Flare rates and prophylaxis use were comparable. At six and twelve months, 62–67% of Group 1 and 83–88% of Group 2 maintained UA targets. Failures were often due to external treatment modifications. No major ULT-related adverse events were observed.

**Conclusion:** Supervised nurse-led gout management effectively achieves UA targets in this “difficult-to-manage” cohort. Maintaining UA targets may benefit from shorter follow-up intervals and enhanced education for patients and healthcare providers.

## Introduction

Gout is among the most prevalent arthritides worldwide, and its incidence continues to rise due to population growth and aging (1). Gouty arthritis causes substantial patient morbidity, including immobility and frequent emergency visits, and carries both direct medical costs and indirect societal costs, such as work absenteeism (2). Beyond the socio-economic impact, gout is an independent cardiovascular risk factor, associated with increased morbidity and mortality (3,4). Despite being a treatable disease, many patients remain undertreated.

Surveys indicate that only 43% of general practitioners correctly identify the target serum urate (UA) levels, and just 32% advocate long-term urate-lowering therapy (ULT) (5).

Rheumatologists are heavily involved when gout becomes symptomatic; however, studies show that only 56% of patients attended by rheumatologists had at least one serum urate measurement (6). In a UK national audit, although rheumatologists initiated or adapted ULT in 76% of indicated patients, only 45% achieved UA <368 µmol/L at 12 months (7), highlighting challenges in achieving optimal control even in specialty care.

Nurse-led management of gout has been shown to outperform usual care in primary care settings, improving UA target attainment, adherence, patient education, satisfaction, flare frequency, and cost efficiency (8–12). Whether a similar model would work in tertiary centers managing polymorbid patients with complex comorbidities and multiple therapeutic restrictions is unclear. These patients often require involvement of multiple disciplines and are more likely to meet criteria for difficult-to-treat gout (13,14), though no validated definition exists. Building on encouraging results from nurse-led outpatient studies (8,9), we implemented a supervised gout outpatient clinic at the University Hospital Bern in 2023 and report our experiences in 69 gout patients, focusing on treatment efficacy and follow-up at six and twelve months.

## Methods

### Study Design

This observational study was conducted at the University Hospital Bern, Switzerland, between May 2023 and January 2026. Patients with confirmed gout who provided informed consent were included. The study was approved by the Cantonal Ethics Committee Bern (No. 2025-01880). Gout diagnosis followed the 2015 ACR classification criteria, including detection of monosodium urate (MSU) crystals in a symptomatic joint, dual-energy CT (DECT), or ultrasound double contour sign (DCS) (15). Patients not fulfilling diagnostic criteria were excluded from ULT.

### Intervention

Initial assessment and all follow-up visits were performed by an Advanced Practice Nurse (APN) in collaboration with a rheumatologist. APN-provided education included disease mechanisms, lifestyle and dietary advice, medication adherence strategies, and delivery of a standardized information booklet. Pharmacological decisions, including ULT initiation and adjustments, flare prophylaxis (colchicine, prednisone, NSAIDs), and SGLT-2 inhibitor use, were made by rheumatologists according to clinical criteria and 2020 ACR guidelines (16).

Allopurinol was the primary ULT, titrated in 50 mg increments for eGFR <30 mL/min/1.73 m^2^ and 100 mg increments for eGFR ≥30 mL/min/1.73 m^2^ until UA targets were reached.

Febuxostat was used for intolerance or prior therapy, starting at 40–80 mg and increased up to 120 mg if needed. Flare prophylaxis followed renal function-adjusted dosing: colchicine 0.5 mg twice daily if eGFR ≥30 mL/min/1.73 m^2^ or once daily if <30, or alternatively low-dose prednisone (5 mg daily) or NSAIDs.

### Cohorts and Treatment Targets

#### Patients were assigned to two predefined UA targets

Group 1 (<300 µmol/L): included patients with tophaceous, erosive, or persistent flares despite prior UA target achievement.

Group 2 (<360 µmol/L): included patients with non-tophaceous, non-erosive gout.

### Follow-Up Procedures

Follow-up visits occurred every 2–4 weeks and included clinical assessment, laboratory testing (CRP, serum urate, creatinine/eGFR, cholesterol, HbA1c), and documentation of BMI, blood pressure, treatment adherence, and flares. UA-targeted ULT adjustments were made at each visit. After achieving UA targets, follow-up visits were scheduled at six and twelve months to assess stability of UA target. UA exceedances prompted ULT adaptation with repeat follow-up in 2–4 weeks. Flares were recorded when fulfilling typical inflammatory patterns (17), such as acute pain, swelling, and allodynia, not explained by trauma or exercise.

### Data Analysis

Continuous variables are presented as mean ± SD, categorical variables as percentages. Group comparisons were performed using Welch t-tests or Fisher’s exact test. A p-value <0.05 was considered statistically significant.

## Results

### Patient Characteristics

Baseline characteristics were comparable between groups (Table 1). Gout was confirmed by MSU, DECT, or DCS in 94% of cases; four patients were diagnosed based on tophi (n=3) or MTP I erosions (n=1). Fifteen patients (22%) were transplanted (14 received immunosuppressants, one undergoing CAR-T therapy) highlighting the complexity of this tertiary cohort.

**Table 1:**
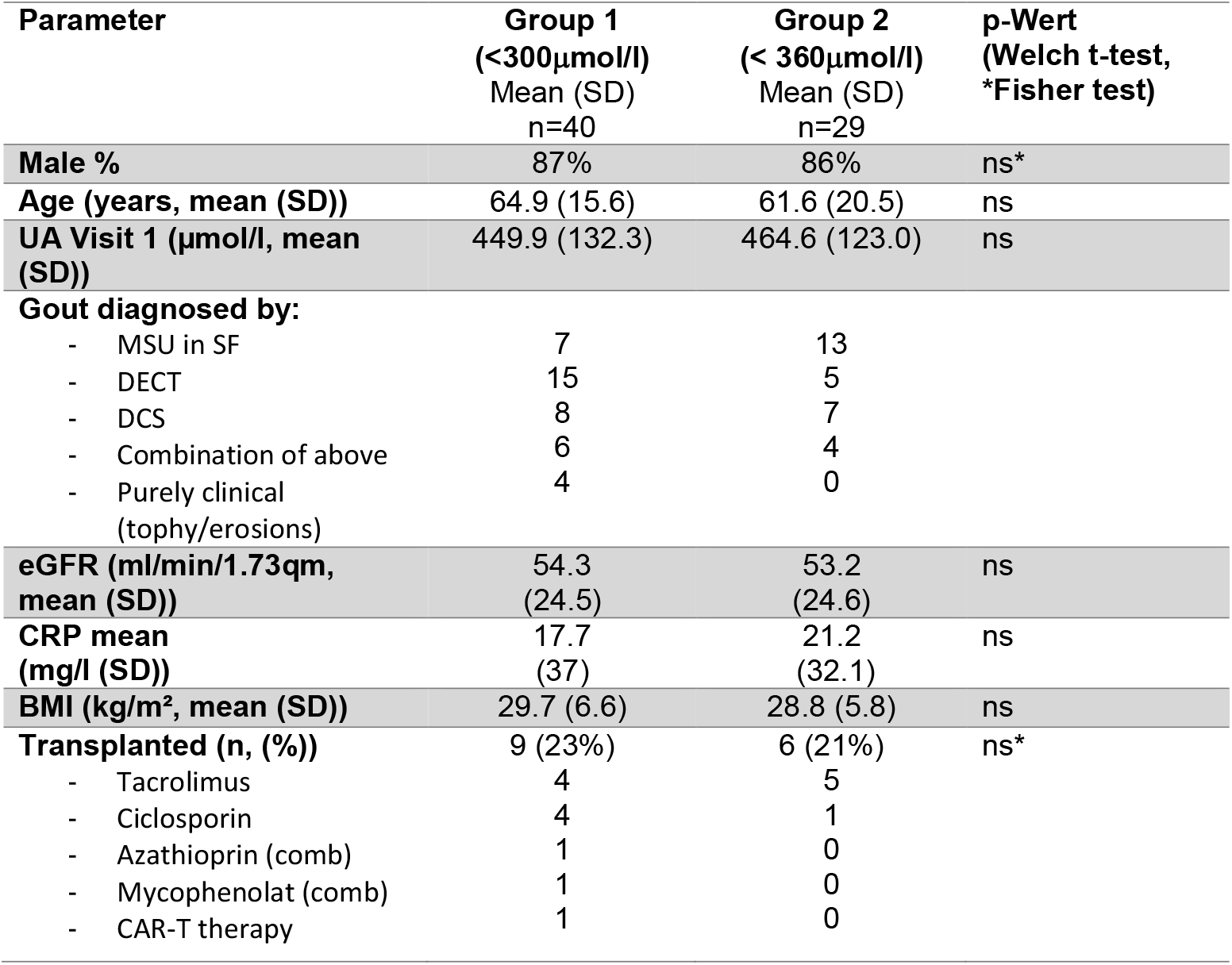
baseline parameters of admitted patients with confirmed gout.

| Parameter | Group 1<br>($<300\mu\text{mol/l}$)<br>Mean (SD)<br>n=40 | Group 2<br>($< 360\mu\text{mol/l}$)<br>Mean (SD)<br>n=29 | p-Wert<br>(Welch t-test,<br>*Fisher test) |
| --- | --- | --- | --- |
| Male % | 87% | 86% | ns* |
| Age (years, mean (SD)) | 64.9 (15.6) | 61.6 (20.5) | ns |
| UA Visit 1 ($\mu\text{mol/l}$, mean (SD)) | 449.9 (132.3) | 464.6 (123.0) | ns |
| **Gout diagnosed by:** |  |  |  |
| - MSU in SF | 7 | 13 |  |
| - DECT | 15 | 5 |  |
| - DCS | 8 | 7 |  |
| - Combination of above | 6 | 4 |  |
| - Purely clinical<br>(tophy/erosions) | 4 | 0 |  |
| eGFR (ml/min/1.73qm, mean (SD)) | 54.3<br>(24.5) | 53.2<br>(24.6) | ns |
| CRP mean<br>(mg/l (SD)) | 17.7<br>(37) | 21.2<br>(32.1) | ns |
| BMI ($\text{kg/m}^2$, mean (SD)) | 29.7 (6.6) | 28.8 (5.8) | ns |
| Transplanted (n, (%)) | 9 (23%) | 6 (21%) | ns* |
| - Tacrolimus | 4 | 5 |  |
| - Ciclosporin | 4 | 1 |  |
| - Azathioprin (comb) | 1 | 0 |  |
| - Mycophenolat (comb) | 1 | 0 |  |
| - CAR-T therapy | 1 | 0 |  |

### Treatment Effectiveness

In Group 1, four patients were within UA target at baseline, and 44 treatment regimens (considering eight re-initiations/adaptions) were implemented in 36 patients. UA <300 µmol/L was achieved in 36/44 regimens (82%), with four patients still undergoing treatment at the time of this manuscript. In Group 2, eight patients were at target at baseline, and 22 regimens (considering one adaption) were conducted among 21 patients, achieving UA <360 µmol/L in 15/22 regimens (68%). Six patients in Group 1 and five in Group 2 were excluded from the analysis due to death, intolerance to therapy or loss to follow-up (Table 2).

**Table 2:** characteristics of treatment regimens achieving UA target.

| Variable | Group 1<br>(<300μmol/l)<br>(n=36) | Group 2<br>(<360μmol/l)<br>(n=15) | Significance<br>(Welch t-test,<br>*Fisher test) |
| --- | --- | --- | --- |
| **Treatment period:** |  |  |  |
| Total number of visits | 122 | 41 | N/A |
| UA mean visit 1 (μmol/l, (SD)) | 441 (124) | 496 (101) | ns |
| with allopurinol | 33 (92%) | 11 (80%) | ns* |
| with febuxostat | 3 (8.3%) | 3 (20%) | ns* |
| Colchicine prophylaxis | 22 (61.1%) | 8 (53.3%) | ns* |
| Prednisolone prophylaxis | 10 (27.8%) | 4 (26.7) | ns* |
| NSAR prophylaxis | 1 (2.8%) | 0 | ns* |
| SGLT2 inhibitors | 10 (27.8%) | 1 (6.7%) | ns* |
| **Until UA target:** |  |  |  |
| Visits until target (mean, (SD)) | 3.4 (1.8) | 2.7 (0.9) | ns |
| Allopurinol dose on target<br>(mg/d, (SD)) | 344 (120) | 179 (84) | **p<0.01** |
| Febuxostat dose on target<br>(mg/d) (n=3 per group) | 80 ± 40 | 53.3 ± 23.1 | N/A |
| Flares total (rate per patient) | 17 (0.47) | 7 (0.47) | ns |
| **Excluded from analysis**<br>**(drop-outs):** |  |  |  |
| - death | 3 | 3 |  |
| - intolerance to treatment | 1 | 1 |  |
| - loss to follow-up | 2 | 1 |  |

**Table 3:** Maintenance of UA target levels at six and twelve months follow-up and presumed reasons for UA target deviation.

|  | Group 1 |  | Group 2 |  |
| --- | --- | --- | --- | --- |
| Follow-up timepoint | 6 months<br>(n=29) | 12 months<br>(n=9) | 6 months<br>(n=12) | 12 months<br>(n=8) |
| **On target** | 62% | 67% | 83% | 88% |
| **Off target** |  |  |  |  |
| - external factors | 7% | 33% | 17% | 0% |
| - non-adherence | 21% | 0% | 0% | 0% |
| - unclear | 10% | 0% | 0% | 12% |

Group 1 required significantly higher allopurinol doses than Group 2 (p<0.01), reflecting lower UA targets and higher prevalence of high-grade renal disease eGFR <30ml/min/1.73sqm and, thus, more patients requiring 50mg increments (n=9 vs. n=5). Flare prophylaxis (colchicine or prednisone) and flare rates were comparable between groups, though one patient in Group 1 accounted for 7/17 flares.

### UA Target Maintenance

At six months, 62% of Group 1 and 83% of Group 2 maintained UA targets; at twelve months, rates were similar. Most UA exceedances in Group 1 were modest (20–30% above target), indicating continued but suboptimal treatment. In 7/17 (41%) of cases, UA target failures were due to treatment adjustments by external physicians, including complete stop of medication, with no imminent adverse events attributable to ULT or prior consultation with us.

## Discussion

Our study demonstrates that a supervised nurse-led gout clinic is effective in achieving UA targets in a complex tertiary cohort. Fifty-eight percent of patients had severe gout, and 15 were transplanted, fulfilling at least one criterion of the previously described “difficult-to-treat gout” (13,14). UA targets were typically reached within 2–4 visits, with allopurinol doses of 200–400 mg in most patients. These findings align with prior primary care studies (8), though our cohort used smaller allopurinol doses (mean 185–347 mg/day vs 460 mg/day), possibly due to smaller increments for low eGFR (50mg increments in those with eGFR <30ml/min/1.73sqm). Severe gout patients required significantly higher allopurinol doses and had slower UA reduction.

Maintenance at six and twelve months was ∼70%, with severe gout patients less likely to remain at target. Interestingly, failure of maintaining UA target levels was due to dose adaptions or even treatment discontinuation by external physicians in almost half of patients. This confirms previous findings from the UK (11) and indicates that both education of patients and attending physicians as well as other health care providers remains an important area to improve interdisciplinary gout management. This may be especially important in multimorbid patients, where more health care providers and an elevated number of communication interfaces exist. Consistently, our findings also suggest that follow-up intervals of six months may be too wide in patients with severe gout in order to maintain UA target levels. New approaches, such as telemedicine (17) and UA self-measurement devices may facilitate UA monitoring (18) and thereby contribute to treatment continuity and better long-term results.

In summary, our data support the feasibility and effectiveness of nurse-led gout management, with outcomes comparable to those reported previously. However, the complexity observed in tertiary-care gout cohorts appears to be driven primarily by multimorbidity and treatment constraints, suggesting a state of difficult-to-manage gout rather than true difficult-to-treat disease, which would imply intrinsic therapeutic resistance.

No major ULT adverse effects were observed. One patient switched from allopurinol to febuxostat due to intolerance. We observed a mortality rate of 11.1% (6/54 treated patients, three in each group) during the 2.5 year observation period. It Is well known that gout patient have an elevated cardiovascular mortality (3) and based on standardized mortality ratios in gout patients (19), an expected mortality rate of 7-8% can be expected during a 2.5 year interval. In a Danish study on nurse-led gout patients revealed a mortality rate of 15% during 2 years (9). We therefore consider the observed mortality rate within the expected range.

Our study has limitations, namely being a single-center observational study without a control arm and showing a highly biased and possibly heterogeneous gout population, limiting generalizability and causal inference. Second, the sample size was modest, increasing susceptibility to individual patient effects. Thirdly, despite following ACR 2020 treatment guidelines external provider interventions may have affected UA target maintenance.

Eventually, the twelve months follow-up period was rather short, limiting conclusions on larger and longer scale.

On a positive note, however, we herein report on a real-world population, without study biases and can widely reproduce the positive findings of larger studies, despite these limitations.

### Key message

Supervised nurse-led gout management at a tertiary center achieved UA targets and adherence in complex-to-manage gout patients comparable to prior studies. Severe gout patients may benefit from shorter follow-up intervals and enhanced education for patients and healthcare providers. Larger multicenter studies are warranted to validate these findings and optimize care for complex gout populations.

## Data Availability

All data produced in the present study are available upon reasonable request to the authors

## Abbreviations

UA: Serum uric acid
ULT: Urate-lowering therapy
GP: General practitioner
APN: Advanced Practice Nurse
MSU: Monosodium urate
DECT: Dual-energy computed tomography
DCS: Double contour sign (ultrasound)
eGFR: Estimated glomerular filtration rate
NSAID: Non-steroidal anti-inflammatory drug
HbA1c: Hemoglobin A1c
CAR-T: Chimeric antigen receptor T-cell therapy
SD: Standard deviation
CRP: C-reactive protein
MTP: Metatarsophalangeal joint

